# Clinical Evaluation of a Digital Biomarker for Joint Swelling in Inflammatory Arthritis based on Automated Quantification of Dorsal Finger Fold Patterns

**DOI:** 10.64898/2026.02.26.26347165

**Authors:** Cinja Koller, Jules Maglione, Marc Blanchard, Arnd Kleyer, Lukas Folle, Jeroen Geurts, Thomas Hügle

## Abstract

**Objective:** To clinically evaluate a digital biomarker, the Finger Fold Index (FFI), derived from the ratio of joint diameter to finger fold surface area in hand photographs, for assessing joint swelling in inflammatory arthritis.

**Methods:** Smartphone hand photographs from two routine care cohorts of patients with rheumatoid (RA) and psoriatic arthritis (PsA) were analyzed using a machine learning pipeline for automated detection and processing of proximal interphalangeal (PIP) joints. The FFI was clinically evaluated by correlation with joint swelling scores (0-3) and DAS28-CRP. A healthy cohort was used to establish FFI reference ranges, which were then compared to the arthritis cohorts.

**Results:** A total of 1275 PIP joint images of 124 arthritis patients and 53 healthy individuals were included. FFI values correlated with swelling scores in the arthritis population with r = 0.443 (95% CI 0.384–0.498). A correlation was observed between the mean FFI and DAS28-CRP dichotomized at 3.2 (r = 0.310, 95% CI 0.123–0.475). FFI values exceeding the healthy reference ranges were associated with swelling (Cramer’s V = 0.400–0.631; *p* < 0.001).

**Conclusion:** FFI values derived from hand photographs showed a significant association with clinical joint swelling and disease activity in RA and PsA patients. Longitudinal studies are needed to assess sensitivity to change and to establish whether this biomarker can be reliably used for remote patient monitoring.

## INTRODUCTION

Remote patient monitoring (RPM) outside medical consultations offers more flexible and effective care in chronic immune-mediated rheumatic diseases (1,2). By facilitating the early recognition of disease flares, or new disease onset in at-risk individuals, RPM can streamline disease management, for example, by helping prioritize in-person consultations or by supporting teleconsultations in remote areas (3,4). In rheumatoid arthritis (RA), the use of digital health applications with patient-reported outcomes (PROs) has been shown to improve disease control rate and cost-effectiveness in real-world settings (5,6). This digital support can be applied and adapted to different disease states and patient pathways such as active or stable disease by detecting flares or treatment responses during treat-to-target or tapering strategies (7). Outcome Measures in Rheumatology Clinical Trials (OMERACT) strongly supports the use of PROs for pain, patient global assessment, physical function, and fatigue (8). However, patient global assessments, often yield higher scores compared to the physician global assessment and are poorly correlated (9,10). This discrepancy is influenced by factors such as fibromyalgia, catastrophizing or other comorbidities that affect the patient’s overall perception of their condition (11). Therefore interpreting PROs can be challenging, and the discordance between physician and patient evaluations complicates clinical decision-making, especially in remote care (12). This also affects other disease activity scores such as DAS28-CRP, where remission is hindered by elevated patient global assessment scores, despite the absence of objective swollen joints (13). Digital biomarkers derived from wearables or behavioral monitoring such as grip strength, movement or sleep analysis have shown potential but lack specificity and are influenced by non-inflammatory factors such as infection, obesity or depression (14).

Joint swelling remains the most objective clinical hallmark of arthritis, typically reflecting synovitis but also occurring with tenosynovitis or dactylitis especially in psoriatic arthritis (PsA). Although ultrasound and magnetic resonance imaging (MRI) provide higher sensitivity for detecting synovitis, their application in remote care is limited by cost, accessibility and operator dependency (15–17). This highlights the need for digital biomarkers that can objectively assess joint swelling in settings suitable for patient self-assessment.

We previously developed a machine learning model for automated analysis of dorsal finger fold patterns of PIP joints on hand photographs to predict clinical joint swelling in RA (18). That initial model captured dorsal finger fold patterns, which were used to train a convolutional neural network (CNN) to classify the images as showing either swollen or non-swollen joints. The number of captured finger folds and the diameter-to-finger-fold-length ratio were quantified manually. In the present study we introduce a refined algorithm pipeline that measures the fold surface area relative to joint diameter on smartphone images, resulting in the finger fold index (FFI). This updated approach provides a more robust and reproducible estimate of PIP joint skin morphology across joints and time points. In this study, we evaluate the FFI as a potential digital biomarker of joint swelling in inflammatory arthritis and explore its association with clinical swelling scores and composite disease activity measures.

## PATIENTS AND METHODS

### Patients and study design

Patients diagnosed with RA or PsA fulfilling the ACR/EULAR 2010 classification criteria (19) and aged ≥18 years were eligible for this study. This prospective study collected smartphone-taken hand images from patients at the University Hospital in Lausanne, Switzerland, and the University Hospital Erlangen, Germany, between 2021 and 2024, alongside clinical data and patient characteristics during regular consultations.

### Ethical considerations

Written informed consent was obtained from all participants, and the study was approved by the Commission Cantonale d’Ethique de la Recherche sur l’Être Humain, Canton de Vaud, Lausanne (CER-VD 2020-00033), Switzerland, and the ethics committee of Friedrich-Alexander-Universität (Dec. 2021/No. 21-422-B), Germany. The study was conducted ethically in accordance with the World Medical Association Declaration of Helsinki.

### Clinical assessment

During consultation the disease activity score (DAS) in 28 joints in combination with C-reactive protein (DAS28-CRP) and clinical swelling in PIP joints were collected. PIP joint swelling was scored binary during consultation and retrospectively by a blinded senior rheumatologist on a scale from 0 to 3 (not swollen, mildly swollen, moderately swollen, severely swollen). Patients with DAS28-CRP values < 3.2 were defined as having low or no disease activity, and patients with a DAS28-CRP ≥ 3.2 were considered as experiencing active disease. Age and sex were collected as demographic variables. No demographic information was available for the German dataset.

### Image acquisition and algorithm-based processing

Photos from Lausanne were taken by an iPhone on a white DINA4 sheet, spreading the fingers with a 20-30 centimeters distance between the hand and the camera. Pictures in Erlangen were taken as described previously by Folle et al. (20). A CNN pipeline was used to process the images. MediaPipe was used for detection of anatomical landmarks of the hands including PIP joints, which were cropped from the full hand image (21). A U-Net model was trained to predict skin fold pixel surface and lengths from a total of 220 images. The joint diameter was measured from the cropped image. A heatmap from the U-Net’s intermediate layers was extracted to visualize fold activations (Figure 1). The FFI was defined as joint diameter / Fold area (=FFI surface) PIP joints 2–4 on both hands.

**Figure 1.**
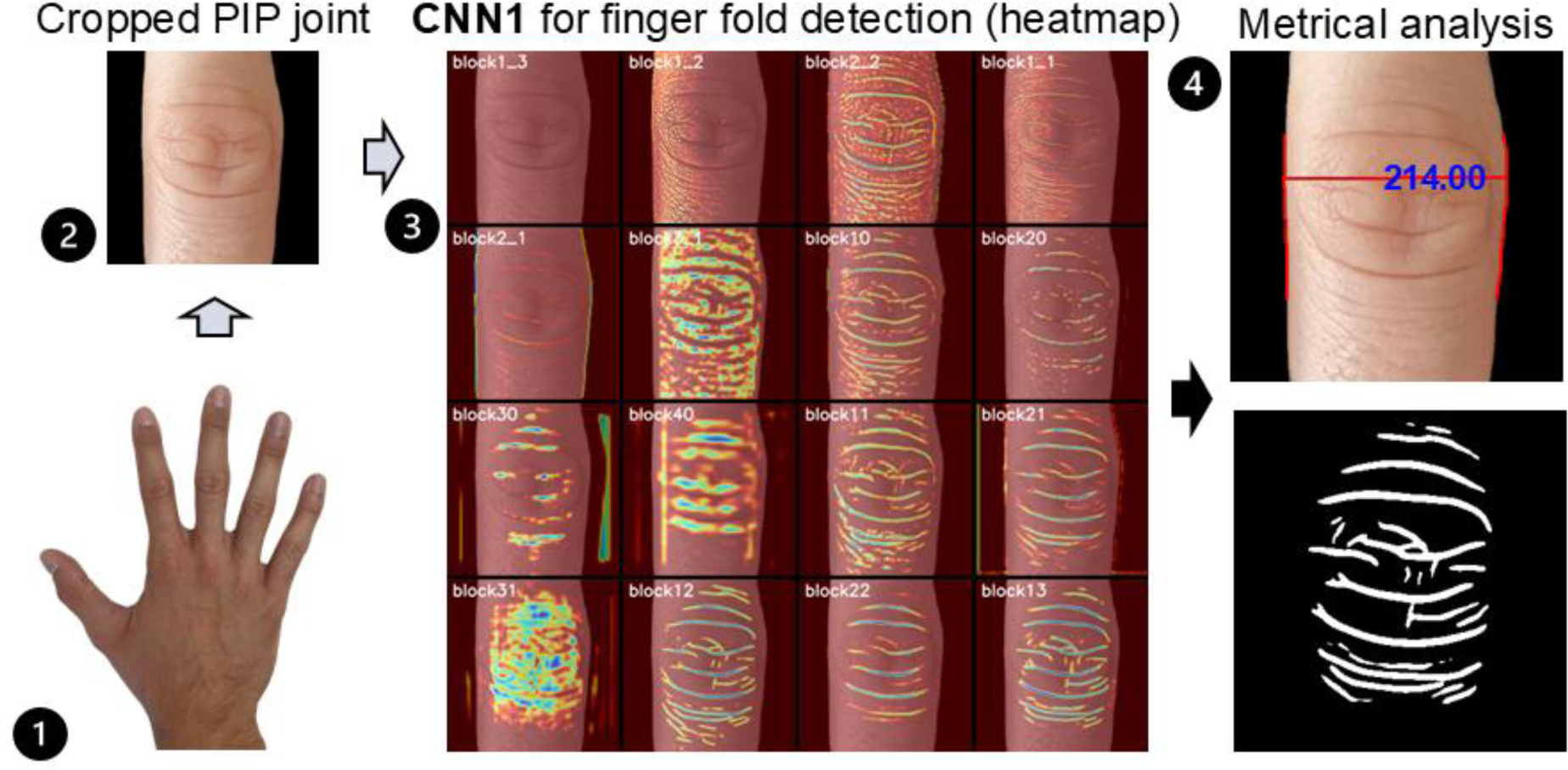
Computer vision and machine learning pipeline. Convolutional Neural Network (CNN), Finger Fold Index (FFI). 1) MediaPipe detects anatomical landmarks on the hand, including the PIP joints. 2) The selected PIP joint is cropped from the full hand image for focused analysis. 3) A U-Net model (CNN1) is trained to predict skin fold pixels. A heatmap is extracted from its intermediate layers to visualize fold activations. The final binary mask indicates the predicted fold locations. 4) The joint diameter is measured from the cropped image. The fold area is quantified by counting the white pixels in the binary mask. Calculation of the FFI = Joint diameter / Fold area. This index quantifies fold density relative to joint size.

### Statistical Analysis

No formal sample size estimation was undertaken given that the main analysis involved correlation testing. To meet general statistical recommendations for robust correlation analysis, data were collected until a minimum of 40 swollen joints per finger was achieved. All eligible participants meeting the inclusion criteria within the study timeframe were included. Missing data were reported, but cases were retained for analysis. Missing values were handled on a variable-by-variable basis, ensuring that available data for other variables within the same case were preserved. We calculated the percentage, mean ± SD for descriptive statistics.

Statistical analyses were conducted separately for RA and PsA and pooled as the arthritis cohort (RA + PsA). We performed spearman’s rank correlation to assess the relationship between swelling (0–3) and the FFI value. In a second analysis, swelling scores were dichotomized (swollen vs. not swollen), and correlations were re-evaluated separately for each finger across the three groups. A Chi-square test was used to examine the association between FFI group membership relative to the norm range and swelling severity within the arthritis cohort. Cramer’s V was calculated to evaluate the strength of the association, with thresholds interpreted as follows: ≤ 0.1 (weak), 0.1–0.3 (moderate), and ≥ 0.3 (strong). The Pearson and spearman’s rank correlation coefficient was used to assess the relationship between the FFI and the DAS28-CRP. *P*-values < 0.05 were considered statistically significant, and 95% confidence interval (CI) were used, except for FFI ranges, where a 99% CI was applied. The CI were not adjusted for multiple comparisons, and therefore the inferences based on them should be interpreted with caution. All analyses were performed with IBM SPSS Statistics version 29.0.2.0 (20) and GraphPad Prism version 10.3.0 (22).

## RESULTS

### Prevalence of joint swelling, distribution of FFI values and patient demographics

We obtained 425 hand photographs from 225 routine care visits across two study sites. PIP joints (digits 2–4) were automatically detected and cropped, yielding 1,275 joint images. Clinical assessment identified swollen joints (score ≥1) in 61.1% (n = 450) of PIP joints in the RA subgroup and 81.6% (n = 186) in the PsA subgroup (Table 1). FFI values were obtained and quantified using the machine learning pipeline (Figure 1). A reference range for each finger (PIP 2-4) on both the left and the right side was established based on data from 53 healthy participants with 103 hand photographs. The upper bound of the 99% CI was used as the cut-off for each finger. The distribution of FFI values, means, and cut-off values for each PIP are summarized in Figure 2. FFI values showed no correlation with age or sex (Supplemental Figure 1-2).

**Figure 2.**
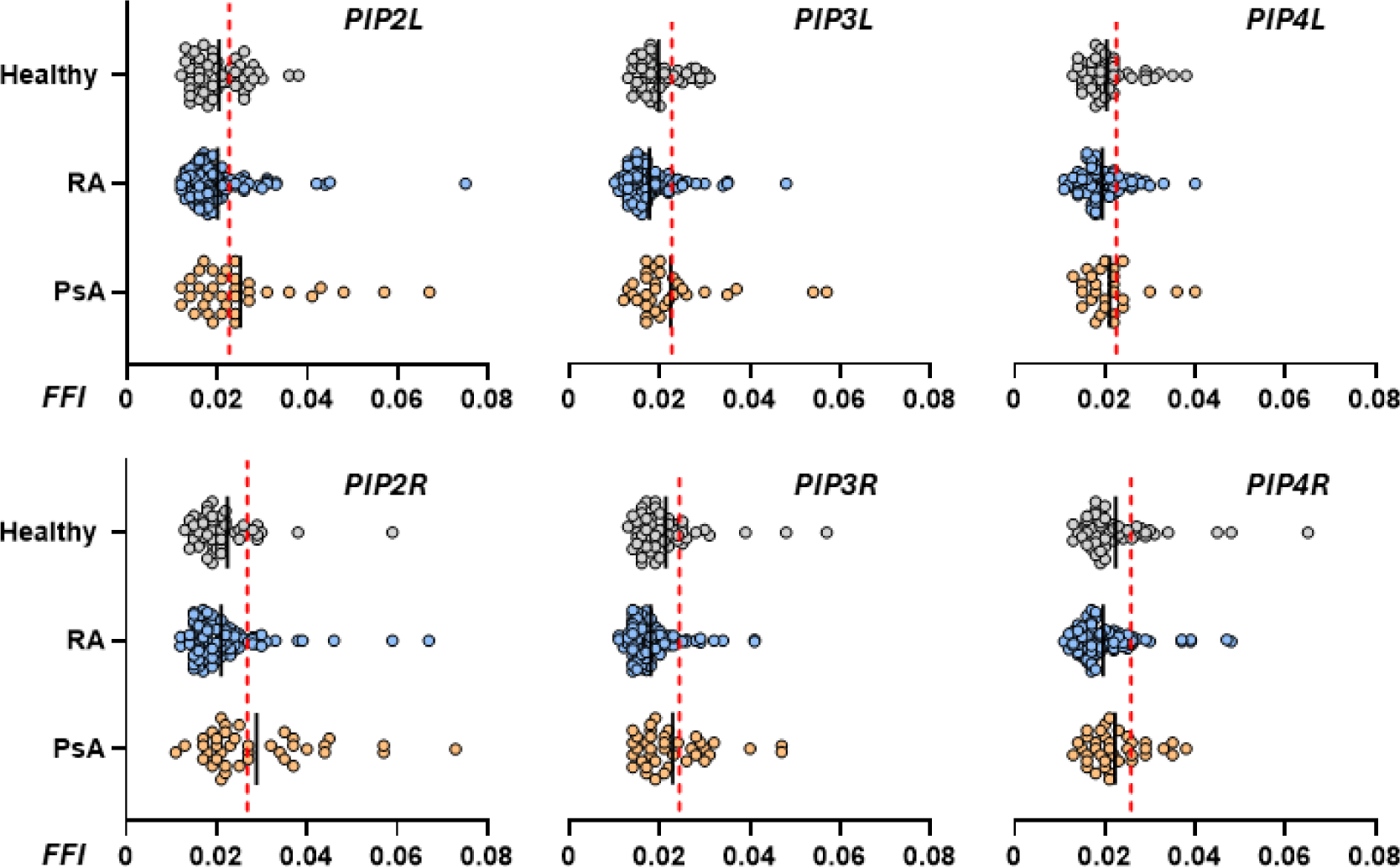
Distribution of FFI values in PIP 2-4 among the healthy and arthritis population. Black Bar = mean, Red dashed line = 99% CI upper limit (reference cut-off).

**Table 1.**
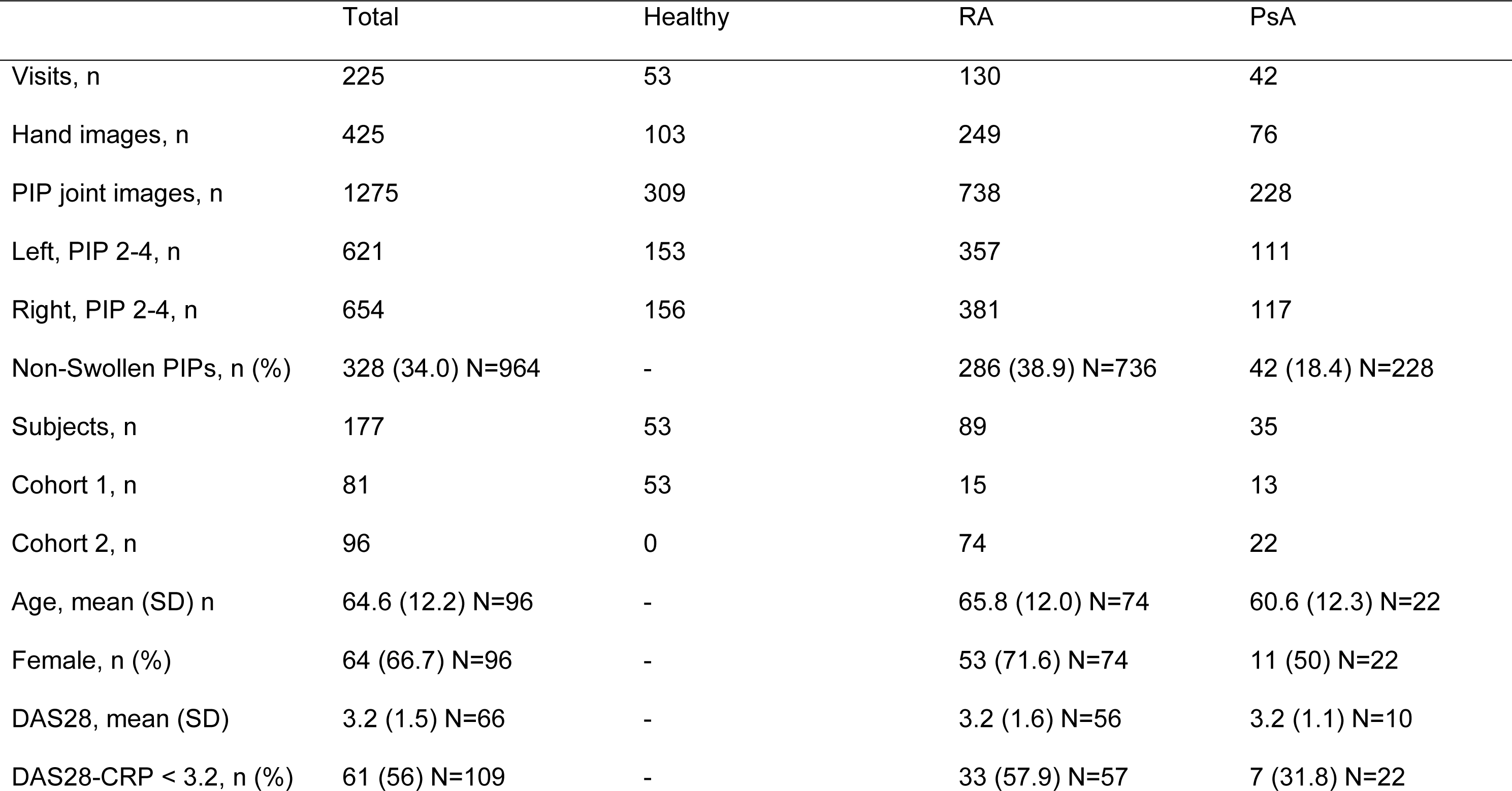
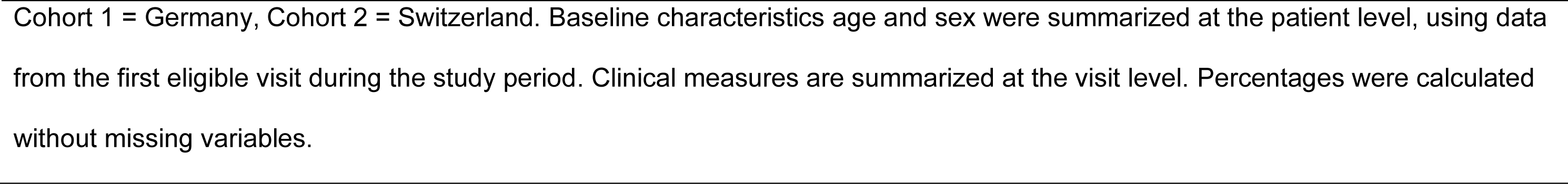
Hand photographs, demographics and clinical characteristics.

|  | Total | Healthy | RA | PsA |
| --- | --- | --- | --- | --- |
| Visits, n | 225 | 53 | 130 | 42 |
| Hand images, n | 425 | 103 | 249 | 76 |
| PIP joint images, n | 1275 | 309 | 738 | 228 |
| Left, PIP 2-4, n | 621 | 153 | 357 | 111 |
| Right, PIP 2-4, n | 654 | 156 | 381 | 117 |
| Non-Swollen PIPs, n (%) | 328 (34.0) N=964 | - | 286 (38.9) N=736 | 42 (18.4) N=228 |
| Subjects, n | 177 | 53 | 89 | 35 |
| Cohort 1, n | 81 | 53 | 15 | 13 |
| Cohort 2, n | 96 | 0 | 74 | 22 |
| Age, mean (SD) n | 64.6 (12.2) N=96 | - | 65.8 (12.0) N=74 | 60.6 (12.3) N=22 |
| Female, n (%) | 64 (66.7) N=96 | - | 53 (71.6) N=74 | 11 (50) N=22 |
| DAS28, mean (SD) | 3.2 (1.5) N=66 | - | 3.2 (1.6) N=56 | 3.2 (1.1) N=10 |
| DAS28-CRP < 3.2, n (%) | 61 (56) N=109 | - | 33 (57.9) N=57 | 7 (31.8) N=22 |

Cohort 1 = Germany, Cohort 2 = Switzerland. Baseline characteristics age and sex were summarized at the patient level, using data from the first eligible visit during the study period. Clinical measures are summarized at the visit level. Percentages were calculated without missing variables.

### FFI correlates with clinical assessment of joint swelling

FFI values showed positive correlations with clinical swelling grades across the second to fourth PIP joints of both hands in the arthritis cohort and within the RA and PsA subgroups (Table 2). Swelling grades from all joints were pooled and correlated with FFI values, yielding r = 0.443 (95% CI 0.384–0.498). In the combined arthritis group on a joint-specific level, correlation coefficients ranged from r = 0.304 to 0.691, indicating weak to strong associations between FFI and swelling severity. Correlations were generally higher when swelling was assessed on the 0–3 scale compared with binary classification. The strongest correlations were observed for the right PIP2 and PIP3 joints in PsA (r = 0.691; 95% CI 0.473 – 0.829 and r = 0.677; 95% CI 0.448 – 0.823) and for the left PIP3 in RA (r = 0.523; 95% CI 0.355 – 0.658).

**Table 2.**
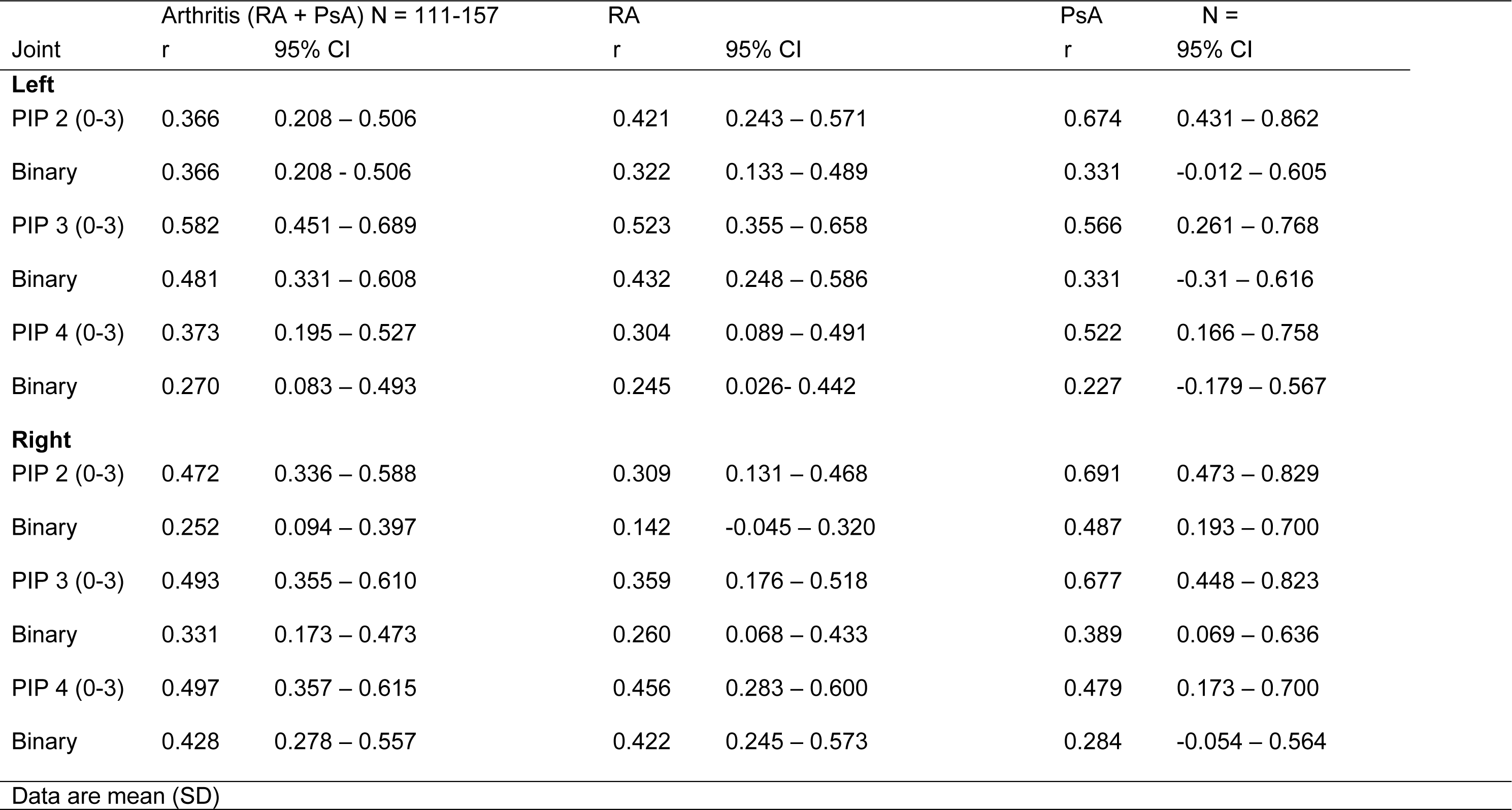
Correlation coefficient (r) and *p* values for swelling (0–3) and the FFI in PIP joints (2–4).

| Joint | Arthritis (RA + PsA) N = 111-157 |  | RA |  | PsA | N = |
| --- | --- | --- | --- | --- | --- | --- |
|  | r | 95% CI | r | 95% CI | r | 95% CI |
| **Left** |  |  |  |  |  |  |
| PIP 2 (0-3) | 0.366 | 0.208 – 0.506 | 0.421 | 0.243 – 0.571 | 0.674 | 0.431 – 0.862 |
| Binary | 0.366 | 0.208 - 0.506 | 0.322 | 0.133 – 0.489 | 0.331 | -0.012 – 0.605 |
| PIP 3 (0-3) | 0.582 | 0.451 – 0.689 | 0.523 | 0.355 – 0.658 | 0.566 | 0.261 – 0.768 |
| Binary | 0.481 | 0.331 – 0.608 | 0.432 | 0.248 – 0.586 | 0.331 | -0.31 – 0.616 |
| PIP 4 (0-3) | 0.373 | 0.195 – 0.527 | 0.304 | 0.089 – 0.491 | 0.522 | 0.166 – 0.758 |
| Binary | 0.270 | 0.083 – 0.493 | 0.245 | 0.026- 0.442 | 0.227 | -0.179 – 0.567 |
| **Right** |  |  |  |  |  |  |
| PIP 2 (0-3) | 0.472 | 0.336 – 0.588 | 0.309 | 0.131 – 0.468 | 0.691 | 0.473 – 0.829 |
| Binary | 0.252 | 0.094 – 0.397 | 0.142 | -0.045 – 0.320 | 0.487 | 0.193 – 0.700 |
| PIP 3 (0-3) | 0.493 | 0.355 – 0.610 | 0.359 | 0.176 – 0.518 | 0.677 | 0.448 – 0.823 |
| Binary | 0.331 | 0.173 – 0.473 | 0.260 | 0.068 – 0.433 | 0.389 | 0.069 – 0.636 |
| PIP 4 (0-3) | 0.497 | 0.357 – 0.615 | 0.456 | 0.283 – 0.600 | 0.479 | 0.173 – 0.700 |
| Binary | 0.428 | 0.278 – 0.557 | 0.422 | 0.245 – 0.573 | 0.284 | -0.054 – 0.564 |
| Data are mean (SD) |  |  |  |  |  |  |

### FFI correlates weakly with disease activity measures

Exploratory analyses examined potential associations between the mean FFI across all PIP joints and DAS28-CRP, a composite measure including joints beyond the fingers and systemic symptoms. Weak correlations were observed with continuous DAS28-CRP (r = 0.152, 95% CI −0.095 – 0.382) and with the binary DAS28-CRP classification (<3.2 vs ≥3.2; r = 0.310, 95% CI 0.123 – 0.475).

### FFI above reference range is associated with severe joint swelling

Cramer’s V showed that FFI values above the reference range were strongly associated with higher swelling severity across all six PIP joints in participants with arthritis (Cramer’s V 0.400–0.631, *p* < 0.001, Table 3). Associations were strongest for right PIP2 (Cramer’s V = 0.631) and PIP3 (Cramer’s = V 0.611) and most pronounced for swelling grade 3.

**Table 3.**
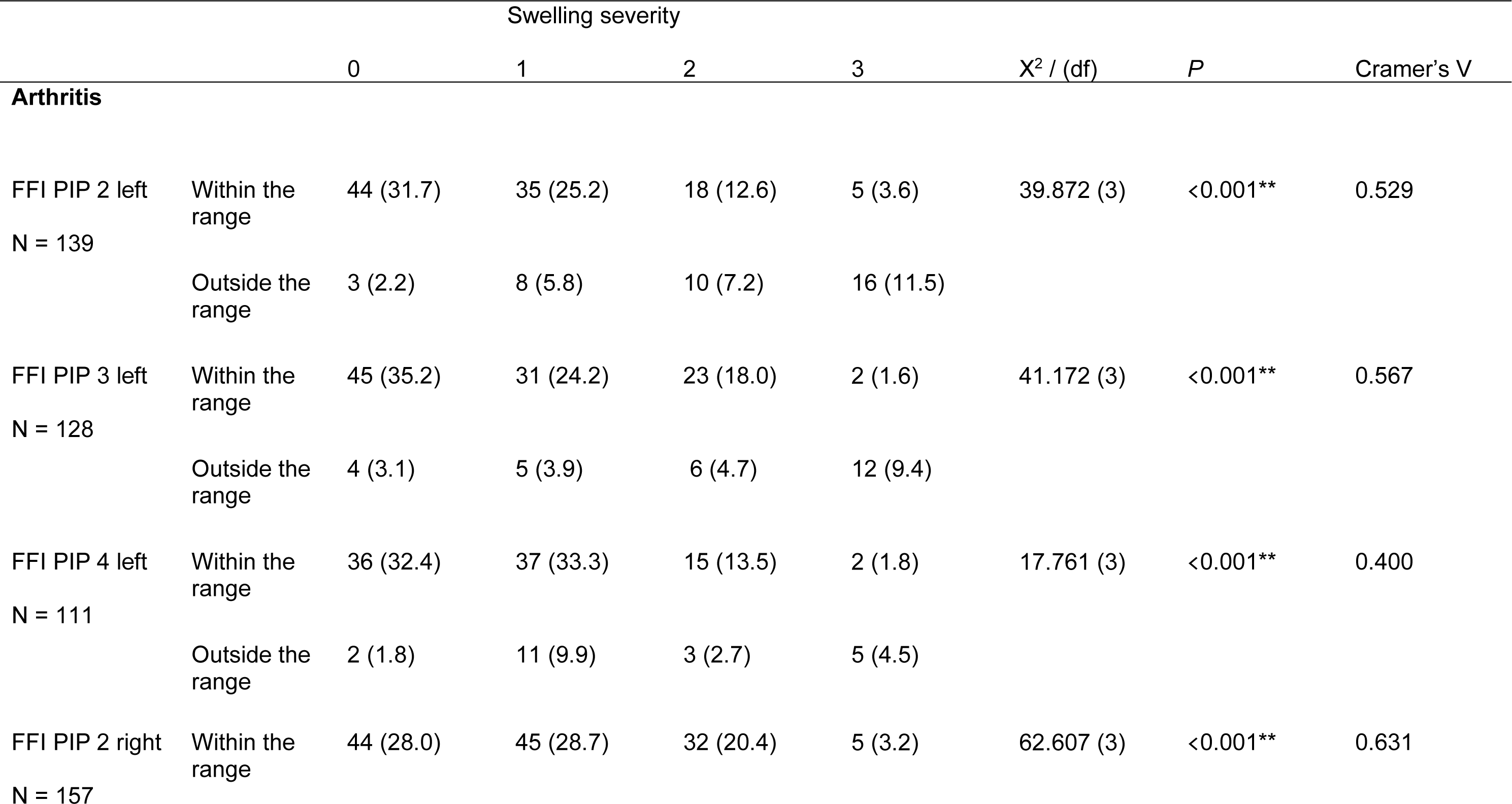
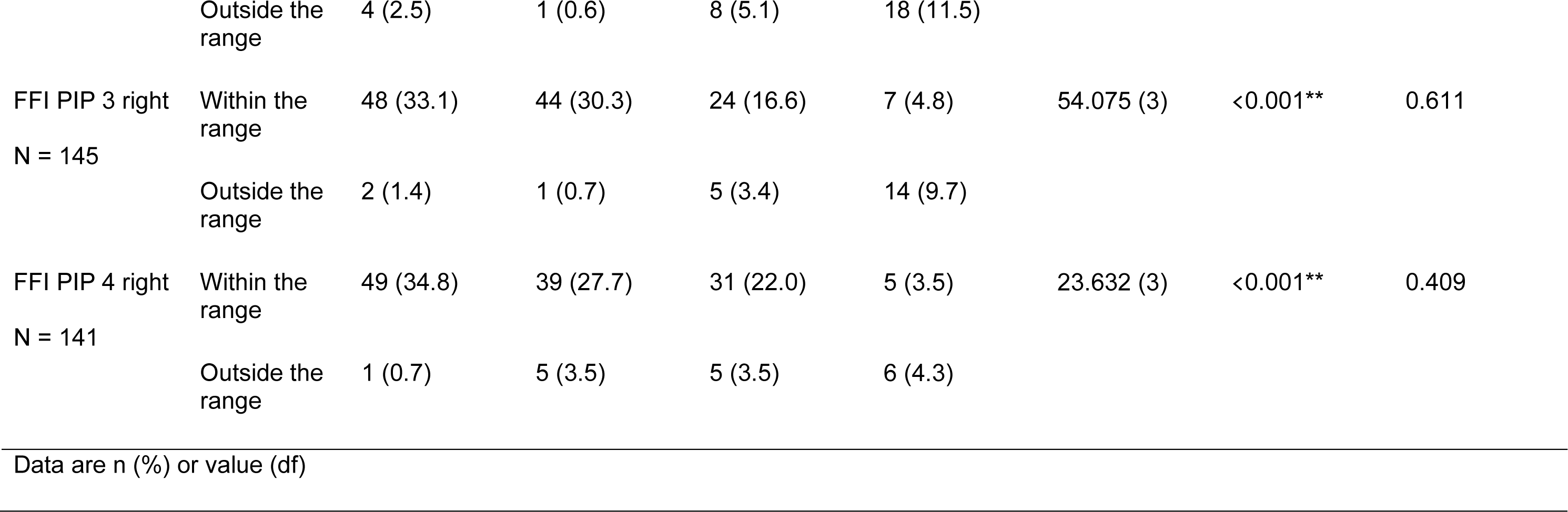
Association of FFI deviation from reference range with PIP joint swelling severity.

## DISCUSSION

A key strength of the FFI is its ability to provide an automated, non-invasive and standardized assessment of PIP joint skin pattern changes, which was strongly associated with clinically detected joint swelling in RA and PsA. Joints with FFI values above the healthy reference range were most likely to exhibit grade 3 swelling, which drove the strongest correlations. The relatively strong associations with swelling grades suggest that changes in FFI may reflect worsening or amelioration of swelling over time. The assessment is rapid and can be performed by patients themselves with proper guidance, making it feasible for repeated measurements in research or integration into RPM, although a numerical value may not be strictly required to detect severe swelling in clinical practice.

Exploratory analyses of FFI and DAS28-CRP scores showed only weak cross-sectional correlations, indicating that FFI is not sensitive enough to replace DAS28 for assessing overall disease activity. In routine care and clinical studies, DAS28 reductions of 0.6–1.2 points are considered moderate improvement and >1.2 points major improvement (23). Longitudinal studies will be necessary to determine whether changes in FFI over time correspond with meaningful clinical improvement. Nonetheless, FFI could be integrated into a remote monitoring platform combining patient-reported outcomes, self- or auto-DAS28, and FFI-based joint swelling assessments, providing complementary information for patient management (24,25).

FFI also showed associations in PsA patients, suggesting it may capture features beyond synovitis, potentially including tenosynovitis or dactylitis. This is a novel observation that warrants further investigation.

This study has several limitations. Healthy reference ranges were derived from a relatively small sample and population characteristics such as age, sex and skin type were not fully represented. Age-related changes in skin tension may influence PIP joint skin morphology in older populations (26). Other potential confounders, such as obesity or fluid retention, were not assessed. Clinical swelling assessment, the ground truth for this study, was performed by a single rheumatologist and imaging modalities such as ultrasound or MRI were not included.

From a technical perspective, false detections, such as misrecognition of rings as anatomical structures, remain a concern. Limitations in image quality, suboptimal hand positioning and difficulty in identifying joints in patients with deformities or active disease underscore the importance of standardized imaging protocols. The model has not yet been validated across different skin tones according to the Fitzpatrick scale (27). Continued algorithm training and improved data acquisition are expected to enhance reliability.

Future improvements to the FFI algorithm could incorporate additional image-based features, such as skin redness, which may provide further information about active inflammation. Combining FFI with tools that assess finger joint motility, such as the MeFISTO technology, could also enhance its ability to capture functional changes in small joints (28). These enhancements, along with longitudinal validation, may improve sensitivity for detecting moderate disease activity changes, flare onset, or treatment response.

In summary, deviations in PIP joint skin patterns captured by the FFI are associated with severe joint swelling in RA and PsA. At present, FFI is primarily useful for identifying joints with pronounced swelling. Its ability to track moderate changes in swelling over time suggests a potential role as a complementary sensitivity-to-change biomarker in remote monitoring platforms, but it does not replace established measures such as DAS28. The rapid and patient-executable nature of the assessment makes it particularly suitable for repeated home-based monitoring. Longitudinal studies and broader validation will be required to determine the clinical utility of FFI, including its integration into patient-centered digital care pathways.

## Data availability

Data can be shared upon justified requests.

## Acknowledgment

We would like to thank participants for their engagement in this study.

