## Supplementary Material for "Clinical Evaluation of a Digital Biomarker for Joint Swelling in Inflammatory Arthritis based on Automated Quantification of Dorsal Finger Fold Patterns"

Figure 1. Distribution of mean FFI surface values per age.

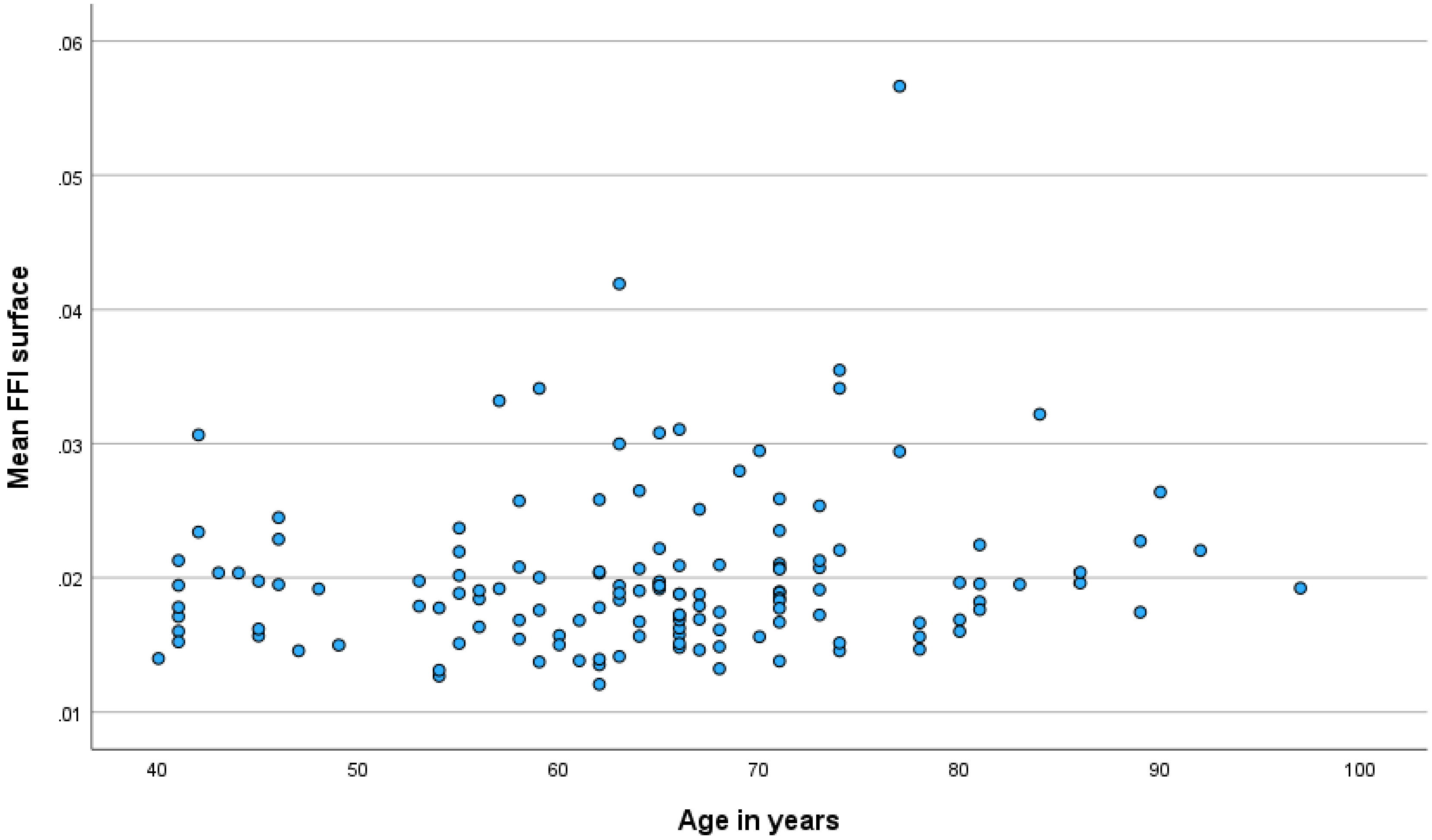

Figure 2. Non-significant difference in mean FFI surface values between male and female subjects.

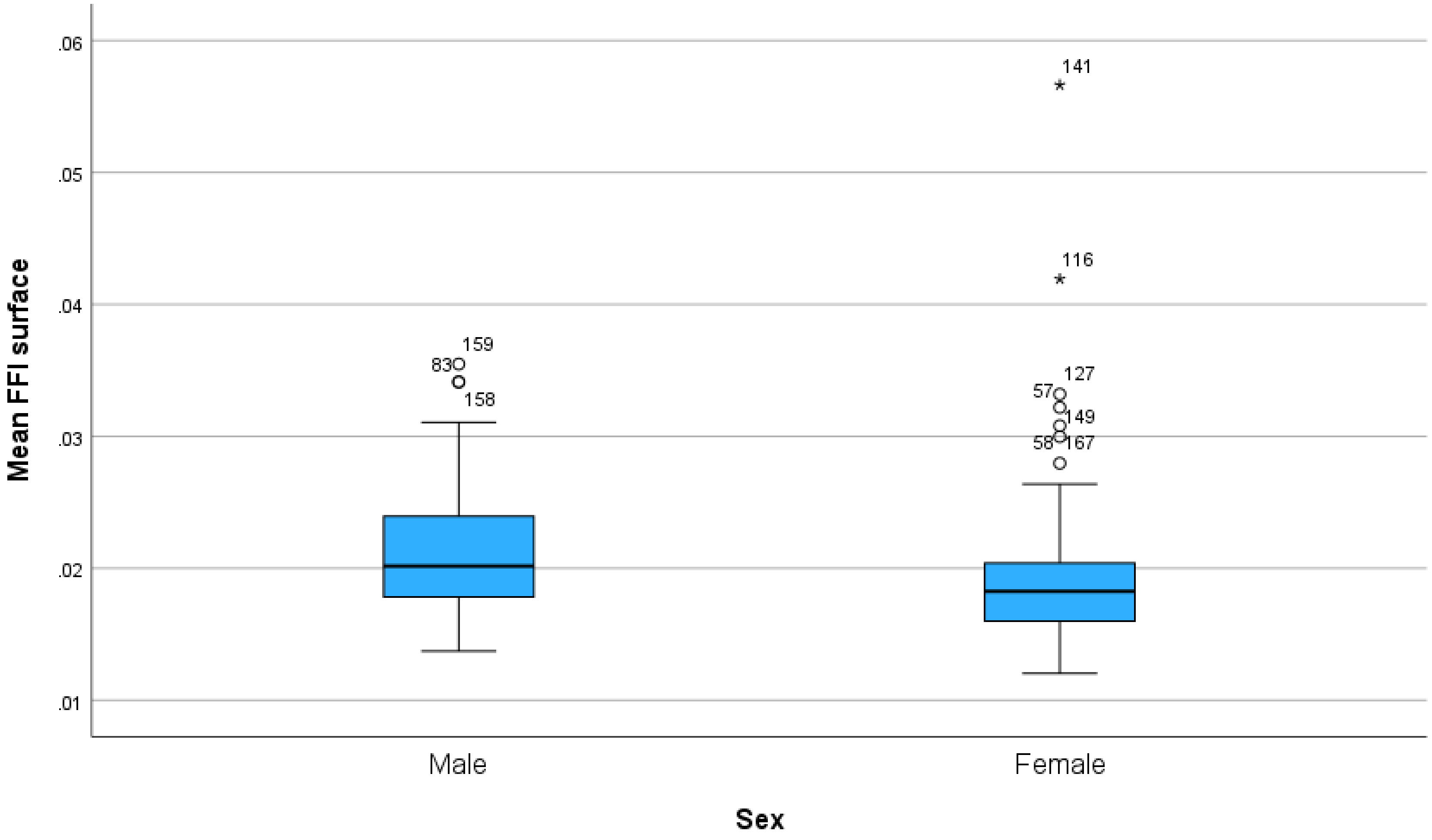
